# Psychosocial distress and health status as risk factors for ten-year major adverse cardiac events and mortality in patients with non-obstructive coronary artery disease

**DOI:** 10.1101/2024.01.11.24301185

**Authors:** Paula M.C. Mommersteeg, Paul Lodder, Wilbert Aarnoudse, Michael Magro, Jos W. Widdershoven

**Affiliations:** CoRPS — Center of Research on Psychology in Somatic diseases, PO box 90153, 5000 LE Tilburg, Tilburg University, the Netherlands; Department of Methodology and Statistics, Tilburg University, the Netherlands; Department of Cardiology, Elisabeth-Tweesteden Hospital, Dr. Deelenlaan 5, 5042 AD Tilburg, The Netherlands

**Keywords:** Non-obstructive coronary artery disease, depression, fatigue, personality, MACE

## Abstract

**Background:** Patients with non-obstructive coronary artery disease (NOCAD) experience psychological distress and are at risk for major adverse cardiac events with mortality (MACE). We examined the risk of psychosocial distress, including Type D personality, depressive symptoms, anxiety, positive mood, hostility, and health status fatigue and disease specific and generic quality of life for MACE in patients with NOCAD.

**Methods:** In the Tweesteden mild stenosis (TWIST) study, 546 patients with NOCAD were followed for 10 years to examine the occurrence of cardiac mortality, a major cardiac event, or non-cardiac mortality in the absence of a cardiac event. Cox proportional hazard models were used to examine the impact of psychosocial distress and health status on the occurrence of MACE while adjusting for age, sex, disease severity, and lifestyle covariates. Sex differences and interactions were explored.

**Results:** In total 19% of the patients (mean age baseline = 61, SD 9 years; 52% women) experienced MACE, with an annualized event rate (AER) of 20 events per 1,000 person-years, and a lower risk for women compared to men. Positive mood (HR 0.97, 95%CI 0.95-1.00), fatigue (HR 1.03, 95%CI 1.00-1.06), and physical limitation (HR 0.99, 95%CI 0.98-1.00) were associated with MACE in adjusted models. No significant interactions between sex and psychosocial factors were present. Depressive symptoms were predictive of MACE, but no longer after adjustment. Neither Type D personality, anxiety, hostility, mental health status, or angina frequency and stability were significantly associated with MACE.

**Conclusions:** In patients with NOCAD fatigue, low positive mood, and a lower physical limitation score were associated with MACE, without marked sex differences. Type D personality, psychosocial factors, and health status were not predictive of adverse outcomes. Reducing psychosocial distress is a valid intervention goal by itself, though it is less likely to affect MACE in patients with NOCAD.

## Introduction

Psychosocial factors have been found to be associated with an increased risk for coronary artery disease incidence, progression, and worse clinical outcomes^1–4^. Examples of such psychosocial risk factors include depressive symptoms, anxiety, hostility, low social support, and low optimism^2^. They also include personality factors such as Type D personality, a combination of high negative affectivity and high social inhibition^5,6^. More so, health status related patient reported outcomes such as fatigue, (vital) exhaustion, health status, and quality of life are not just valuable outcome indicators by itself, but have been associated with adverse clinical outcomes in cardiovascular disease patients^7,8^.

Differences between women and men emerge when examining the risk of psychosocial factors for adverse clinical outcomes in patients with established ischemic heart disease^3^, with men showing a higher risk for adverse outcomes attributable to psychosocial factors compared to women. However, most studies on adverse outcomes are done in patients with *obstructive* ischemic heart disease (IHD), having angina pectoris or an acute myocardial infarction, or who have undergone a percutaneous coronary intervention (PCI), or coronary artery bypass graft surgery (CABG). Studies in patients with non-obstructive IHD, which is more prevalent in women^9^, remain underrepresented.

Having a clinical profile of signs and symptoms of ischemia but without coronary obstructions has been rephrased “open artery ischemia” (OAI)^10^, which can include the presence of angina or ischemia with no obstructive coronary arteries (ANOCA/INOCA). In the present study, the majority of patients meet the ANOCA/INOCA definition, though patients were also included who were referred because of a high familial risk or other risk factors, without current signs and symptoms of either angina or ischemia^11,12^.

In the present study we describe MACE and mortality (referred to as MACE) as a combination of cardiac mortality, major adverse cardiac events and, when absent, noncardiac mortality, in patients with NOCAD across a follow-up time of ten years. The primary goal is to examine psychosocial factors associated with MACE. We hypothesize that having adverse psychosocial factors; Type D personality, depressive symptoms, anxiety, low positive mood, hostility, and worse fatigue, mental, physical, and disease specific health status is associated with a higher risk for MACE. We expect this association both before and after adjustment for disease severity, and adverse lifestyle factors such as being overweight, and being physically active. In secondary analyses we explore the interaction with sex and sex-stratified risk of psychosocial factors for MACE. Subgroup analyses of ANOCA and INOCA for MACE will be examined, as well as MACE excluding cases with noncardiac mortality.

## Methods

### Participants and procedure

The present study is part of the TweeSteden Mild Stenosis study (TWIST), an observational cohort to examine psychosocial risk factors for outcomes in patients with non-obstructive coronary artery disease ^11^(ClinicalTrials.gov Id: NCT01788241). All participants who received a computed tomography (CT scan) or coronary angiography (CAG) were screened between January 2009 and March 2013, and eligible patients received information on the study. Inclusion criteria were having a CT scan with either a coronary artery calcium score ≥ lowest 10th percentile with mild atherosclerotic plaque, excluding those eligible for a subsequent CAG. Separately, all patients undergoing a CAG were examined for having visible wall irregularities without obstructive coronary occlusion (between 10-60% occlusion of coronary arteries). Exclusion criteria were having a history of obstructive coronary heart disease. In total 547 patients provided written informed consent. One person was additionally excluded. Coded paper questionnaires on sociodemographic, lifestyle, and psychological complaints were sent and received by postal mail at baseline, after 12, and 24 months. Electronic hospital record information was used for indicators of disease severity, medication, and comorbid conditions. The TWIST study received ethical approval in 2009 (METC Brabant, Protocol number: NL22258.008.08), with an update for screening for outcomes in 2021-2022 (ETZ: L1160.2020 > LP.2008.227 TWIST follow-up). Study data are stored in accordance with and conforming to good clinical practice and data protection guidelines.

### Primary outcomes

Electronic hospital records were screened for outcomes by trained research assistants (LV, AB, ABE) between October 2021 and February 2022. Outcomes had previously been examined between October 2014 and April 2015^13^. New and previous events were double checked. Raw cardiovascular events with dates and information on the cause of the event at follow up were reported in the local Electronic Data Capture management system (ETZ Research Manager). Additional checks and interpretation of unclear outcomes were done. Coded variables were exported and categorized into primary and secondary outcomes.

The primary outcome was the occurrence of a major adverse cardiac event with mortality (referred to as MACE) during follow-up, including: cardiovascular mortality, cardiac events (PCI, CABG, (N)STEMI or cardiac arrest, heart failure), or non-cardiac mortality. The date of each event was coded. In case of multiple events, hierarchical choices were made; cardiovascular mortality was chosen over a cardiac event, and any cardiac event was chosen over non-cardiac mortality. For descriptive purpose, mortality causes were further subcategorized^14^. Time between the index coronary angiography or CT-scan at inclusion and MACE or censored date was calculated in years. In the absence of an event, patients were censored at the final screening date (February 3, 2022). The censoring date of patients lost to follow-up was set at 12 months after the last moment of contact^15^.

In the Clinicaltrials.gov preregistration done in 2009 we previously defined MACE as ‘the occurence of a recurrent coronary angiography, emergency hospitalization (for cardiac reasons), myocardial infarction, percutaneous coronary intervention (PCI) or coronary artery bypass graft surgery (CABG), mortality (cardiac/noncardiac)’. We choose to deviate from this preregistered definition due to having previously reported on ED visit or repeat testing^13^, to be able to distinguish between cardiac and non-cardiac mortality in the process of disease development, and the previous absence of heart failure as a valid disease progression indicator. In our opinion the current MACE definition better reflects cardiac disease progression, extended with non-cardiac mortality.

### Psychosocial distress

Well validated baseline questionnaires were used^12^. Type D personality was measured using the DS14^5^, measuring negative affectivity (NA) and social inhibition (SI), each with seven items on a five point Likert scale. Individuals were considered to have a Type D personality when they scored 10 or higher on both the NA and SI total score. For the analyses the NA and SI total scores were transformed into z-scores and multiplied to obtain their interaction Z(NA)*Z(SI). Both the z-scores and their interaction were included together as predictors in outcome analysis, with the interaction term testing the Type D personality effect^6^.

Depressive symptoms were assessed with the Beck Depression Inventory (BDI), using a sumscore of the 21 items on a 0-3 range, as well as with the cognitive-affective and somatic subscales (examined together). Separately, the 14 item, 0-3 range hospital anxiety and depression scale (HADS) was used to measure depressive symptoms (HADS-D) and anxiety (HADS-A).

Hostility was operationalized using the Cook-Medley Hostility scale (CMH), containing 27 items on a true/false scale^16^. The sumscore was used as a measure of hostility, with a higher score indicating a more hostile personality style. Positive affect was measured with 10 items on a 0-4 Likert scale from the Global Mood Scale [GMS-PA], with a higher sum score indicating a more positive mood.

### Health status and quality of life

Fatigue was measured using the 10-item fatigue assessment scale [FAS], with a higher score indicating more fatigue. A modified version derived from the Seattle Angina Questionnaire was used to measure disease-specific heart-related complaints in five dimensions^11,17^. A higher score indicates better health and fewer complaints, as well as the generic Short Form 12, derived from the SF36 was used to evaluate physical (physical component summary (PCS)), and mental health (mental component summary (MCS)).

### Descriptive and potential confounding factors

#### Sociodemographic variables

included age, sex, having received at least college education, and having a partner.

#### Disease severity indicators

The cohort was further divided into ANOCA or INOCA^10^ when being referred for (atypical) chest pain or angina pectoris symptoms (ANOCA), or whether additional tests (ECG, ergometry, SPECT scan) indicated (suspect) ischemia (INOCA), versus the patients receiving a diagnostic CAG or CT scan due to having a high familial risk or risk factors (other).

Disease severity was also represented by being included via CAG versus CT scan as per selection procedure, and the latter group more often included people with high familiar risk without clear coronary complaints.

#### Lifestyle factors

included body mass index (BMI), currently smoking (versus former or nonsmoking), any alcohol use versus none, being physically active versus moderate or nonactive. Main categories of *cardiac medication use* were reported, summarized by their Anatomical Therapeutic Chemical (ATC) code, as well as the prevalence of *comorbid conditions*^13^.

### Statistical analysis

Descriptive statistics stratified by presence or absence of MACE were reported and compared between these two groups using Chi-square test for categorical variables, and One-Way ANOVA for continuous scores or a non-parametric equivalent. The annualized event rate (AER) for MACE was calculated dividing the number of MACE events by the sum of total person-years of follow-up (either MACE or censored)*1000, expressed per 1,000 person-years. For comparison reasons, the AER was divided by the average follow-up time, named the ‘Annual MACE rate’^18^.

Cox proportional hazard analyses were performed using MACE as the primary outcome, and time since the baseline CAG or CT in years. An explorative model with sex and age was examined, with sex depicted in a Kaplan-Meier curve. The psychosocial and health status variables of interest were each entered *separately* in an analysis, adjusted for age and sex. In the complete adjusted model the CAG versus CT group and hypertension were added as indicators of disease severity to age and sex, and indicators of lifestyle (BMI and being physically active) since these tend to covary with psychosocial factors^19^.

Sensitivity analyses were done stratified by sex, as well as the interaction of sex-by-psychosocial factor adjusted for age^20^. The Type D personality continuous score was explored, by adding the quadratic negative affectivity and social inhibition terms^21^. Sensitivity analyses for MACE were run with ANOCA and INOCA (reference ‘other’) group as an indicator of disease severity, adjusted for age and sex, as well as excluding non-cardiac mortality cases.

## Results

In total 104 of the 546 patients (19%) with nonobstructive IHD had a major adverse cardiovascular event (MACE) at follow-up, and 81% had cardiac event free survival (81%, n=442). MACE included cardiac mortality (13%, n=15), PCI (32%, n=33), CABG (7%, n=7), (N)STEMI or cardiac arrest (7%, n=7), heart failure (4%, n=4), or non-cardiac mortality (37%, n=38). Time since inclusion was on average 9.5 years (SD = 2.9 years), with a median of 10.3 years (range 0.2-13.2 years). The annualized event rate (AER) for MACE was 20.1 events per 1,000 person-years, or 2.0% (104 MACE events/9.5 years/546 patients). In total 15 people died of cardiovascular causes (heart failure, acute MI, sudden cardiac death, stroke, cardiovascular hemorrhage). Non-cardiac mortality (n=38; excluding four cases who had a cardiac event preceding non-cardiac mortality) was due to malignancies (n=14), pulmonary (n=5), neurological (n=5), trauma (n=4), renal (n=2), COVID (n=2), other hemorrhage causes or other noncardiovascular (n=3), or undetermined (n=3). The AER for all-cause mortality was 11 per 1,000 person years, or 1.1% annually. Patients who had a MACE at follow-up were older and less often higher educated (Table 1), though without significant sex differences (17% (n=47) women; 22% (n=57) men, X^2^ = 2.4, p=.122). The MACE group more often was included via CAG screening (90%) rather than via a CT scan (65%). In total 82% of the patients meet the description for either INOCA (44%) or ANOCA (38%), but no significant differences were observed between the INOCA, ANOCA, and ‘other’ groups for MACE. Patients with MACE did not have significantly different lifestyle factors. Cardiac medication use was not different between the groups, except for more Coumarin derivatives use in the MACE group, but the prevalence was low (7% versus 2%). Hypertension was more prevalent in the MACE group (90% versus 81% in the non-event group).

**Table 1.**
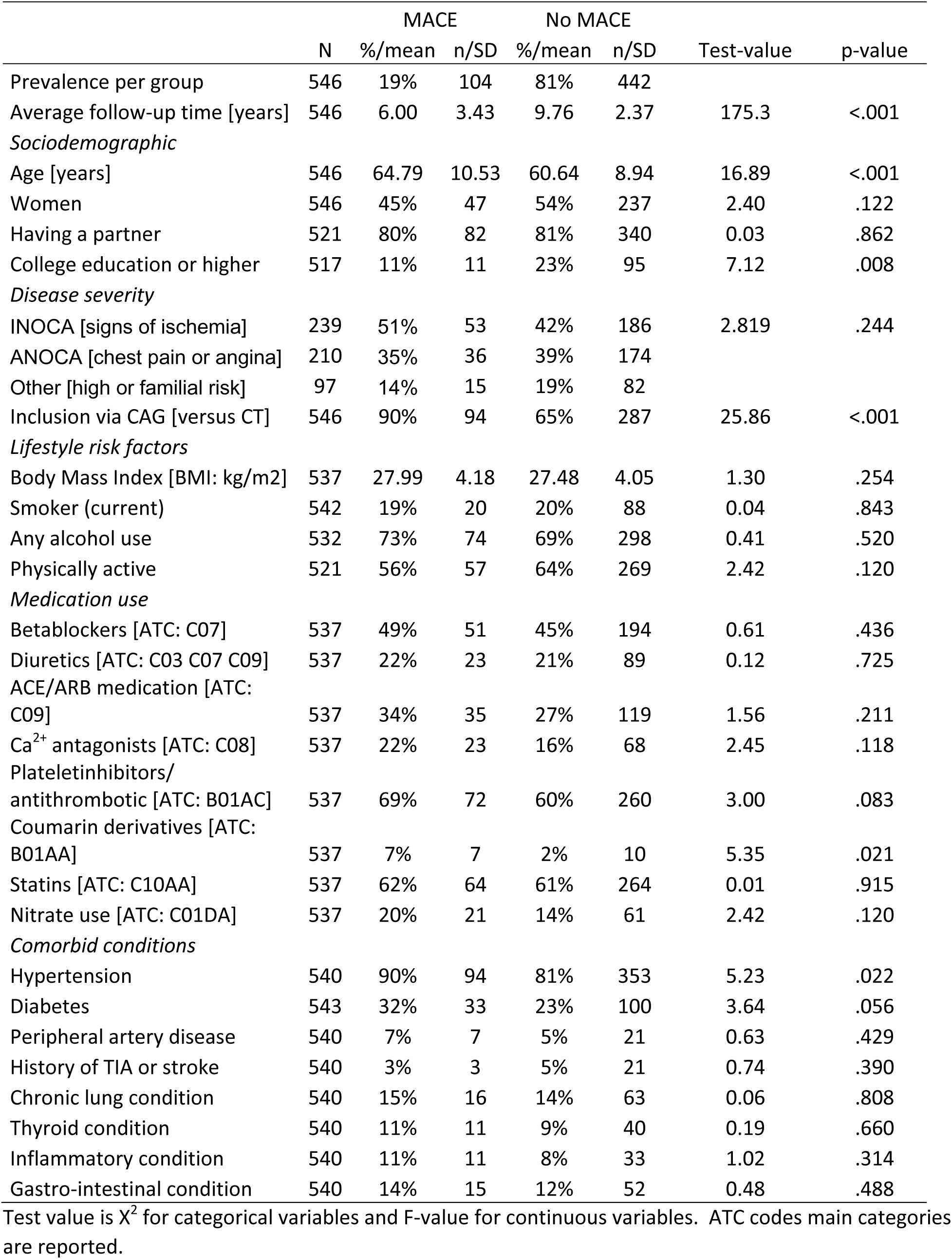
Descriptive factors in patients with NOAD stratified by presence or absence of MACE.

Psychosocial factors and health status scores stratified for MACE were reported in Table 2. Total depressive symptoms as measured by either the BDI, BDI somatic subscale, and HADS were significantly higher in the MACE group, whereas positive mood was significantly lower. No significant differences were observed for Type D personality or its subscales negative affectivity or social inhibition, BDI using cut-off values, anxiety, or hostility. Health status variables fatigue was significantly higher, physical component summary score and the physical limitations score significantly lower in the MACE compared to the group without MACE. No differences were observed for the mental component, angina frequency and stability, treatment satisfaction, or disease perception.

**Table 2.** Psychosocial factors and health status stratified by major adverse cardiovascular events at follow-up.

|  | N | MACE |  | No MACE |  | Test-value | p-value |
| --- | --- | --- | --- | --- | --- | --- | --- |
|  |  | %/mean | n/SD | %/mean | n/SD |  |  |
| Overall prevalence | 521 | 20% | 102 | 80% | 419 |  |  |
| *Psychosocial distress* |  |  |  |  |  |  |  |
| Type D personality [DS14] | 517 | 37% | 36 | 28% | 119 | 2.63 | .105 |
| Negative Affectivity | 516 | 10.15 | 6.92 | 9.53 | 6.14 | 0.78 | .379 |
| Social inhibition | 515 | 9.85 | 6.17 | 9.00 | 6.05 | 1.53 | .217 |
| BDI depressive symptoms [BDI total] | 520 | 10.80 | 8.09 | 9.07 | 7.40 | 4.26 | .039 |
| Cognitive-affective component [BDI cogn] | 520 | 4.80 | 5.22 | 3.97 | 4.91 | 2.20 | .139 |
| Somatic component [BDI som] | 520 | 6.00 | 3.67 | 5.11 | 3.21 | 5.85 | .016 |
| HADS depressive symptoms [HADS-D] | 521 | 6.02 | 4.22 | 4.85 | 3.93 | 7.08 | .008 |
| HADS anxiety [HADS-A] | 521 | 6.75 | 4.63 | 6.15 | 4.13 | 1.67 | .197 |
| Positive mood [GMS-PA] | 513 | 20.08 | 8.75 | 22.69 | 8.21 | 7.79 | .005 |
| Hostility [CMH] | 511 | 12.59 | 5.06 | 11.68 | 5.12 | 2.49 | .115 |
| *Health status and Quality of life* |  |  |  |  |  |  |  |
| Fatigue [FAS] | 520 | 24.47 | 7.45 | 22.56 | 6.81 | 6.15 | .013 |
| Physical Limitations [msAQ] | 500 | 66.94 | 24.56 | 76.90 | 21.00 | 16.55 | <.001 |
| Angina Stability [msAQ] | 515 | 56.31 | 22.68 | 57.03 | 21.47 | 0.09 | .767 |
| Angina Frequency [msAQ] | 546 | 87.50 | 17.67 | 89.68 | 15.79 | 1.54 | .216 |
| Treatment satisfaction [msAQ] | 514 | 80.97 | 18.92 | 82.63 | 18.83 | 0.61 | .434 |
| Disease Perception/ QoL [msAQ] | 509 | 69.74 | 20.24 | 73.76 | 19.21 | 3.42 | .065 |
| *SF36 derived PCS and MCS* |  |  |  |  |  |  |  |
| Physical Component [PCS] | 521 | 41.86 | 10.94 | 44.56 | 10.49 | 5.26 | .022 |
| Mental Component [MCS] | 521 | 42.82 | 12.78 | 44.41 | 11.64 | 1.45 | .229 |

Test value is $\chi^2$ for categorical variables and One-Way ANOVA for continuous variables.

**Table 3.** Cox proportional hazard of psychosocial factors for MACE in patients with NOCAD.

|  | Age and sex<br>adjusted |  | Complete<br>adjusted |  |
| --- | --- | --- | --- | --- |
|  | HR (95%CI) | p-value | HR (95%CI) | p-value |
| *Psychosocial distress* |  |  |  |  |
| Negative Affectivity [z(NA)] * | 1.03 (0.82-1.29) | .814 | 0.99 (0.79-1.24) | .921 |
| Social inhibition [z(SI)] * | 1.15 (0.91-1.44) | .244 | 1.10 (0.88-1.39) | .403 |
| Interaction NA*SI [z(NA)*z(SI)] * | 0.92 (0.74-1.14) | .459 | 0.92 (0.75-1.14) | .448 |
| Type D personality [DS14; dich] | 1.37 (0.91-2.07) | .133 | 1.22 (0.80-1.85) | .353 |
| Depressive symptoms [BDI total] | **1.03 (1.01-1.06)** | **.012** | 1.02 (1.00-1.05) | .104 |
| Cognitive-affective component † | 1.01 (0.96-1.06) | .743 | 1.00 (0.95-1.05) | .936 |
| Somatic component † | 1.07 (0.99-1.15) | .074 | 1.05 (0.97-1.14) | .192 |
| Depressive symptoms [HADS-D] | **1.06 (1.02-1.11)** | **.008** | 1.04 (0.99-1.10) | .087 |
| Anxiety [HADS-A] | 1.04 (0.99-1.08) | .117 | 1.02 (0.97-1.07) | .418 |
| Positive mood [GMS-PA] | **0.96 (0.94-0.99)** | **.002** | **0.97 (0.95-1.00)** | **.048** |
| Hostility [CMH] | 1.02 (0.98-1.06) | .436 | 1.00 (0.97-1.04) | .861 |
| *Health status and Quality of life* |  |  |  |  |
| Fatigue [FAS] | **1.05 (1.02-1.07)** | **.001** | **1.03 (1.00-1.06)** | **.029** |
| Physical limitation [mSAQ] | **0.98 (0.97-0.99)** | **<.001** | **0.99 (0.98-1.00)** | **.021** |
| Angina stability [mSAQ] | 1.00 (0.99-1.01) | .616 | 1.00 (0.99-1.01) | .461 |
| Angina Frequency [mSAQ] | 0.99 (0.98-1.00) | .243 | 1.00 (0.99-1.01) | .871 |
| Treatment Satisfaction [mSAQ] | 0.99 (0.98-1.00) | .229 | 1.00 (0.99-1.01) | .388 |
| Disease perception/ Quality of life [mSAQ] | **0.99 (0.98-1.00)** | **.012** | 0.99 (0.98-1.00) | .096 |
| *SF36 derived PCS and MCS* |  |  |  |  |
| Physical Component [PCS] | **0.98 (0.96-0.99)** | **.009** | 0.99 (0.97-1.01) | .147 |
| Mental Component [MCS] | 0.99 (0.97-1.00) | .130 | 1.00 (0.98-1.01) | .577 |
| The complete adjusted model included sex, age, disease severity (CAG versus CT group, hypertension), and lifestyle factors (BMI and being physically active) |  |  |  |  |

\* Continuous Type D personality variables were combined
† Subscales of BDI; somatic and cognitive/affective component were combined

When examining Cox proportional hazard ratio’s for sex and age, women had a significantly lower risk for MACE than men (HR 0.62, 95%CI 0.42-0.92, p= .016), and age was related to a higher risk for MACE (HR 1.05, 95%CI 1.03-1.07, p<.001). The sex stratified risk is depicted in the Kaplan-Meier curve in Figure 1. Next, Cox regression models were used to investigate if the occurrence of MACE throughout follow-up could be predicted by the psychosocial factors. In the models adjusted for age and sex, depressive symptoms according to the BDI or HADS were related to an increased risk for MACE, whereas positive mood was protective. No significant elevated risk for MACE was observed for Type D personality, anxiety, or hostility. Patient reported health status showed that more fatigue was related to a higher risk for MACE. Higher scores on physical limitation or physical component, and better disease perception (quality of life) was related to a *lower* MACE risk. Adjusting the models for inclusion via CAG, hypertension, and lifestyle factors BMI and being physical active rendered the risk for depressive symptoms no longer significant. Positive mood (HR 0.97, 95%CI 0.95-1.00), fatigue (HR 1.03, 95%CI 1.00-1.06), and physical limitation (HR 0.99, 95%CI 0.98-1.00) remained significantly associated with MACE. In the final model women had a significantly lower risk for MACE compared to men (HR 0.60, 95%CI 0.40-0.89), whereas being older (HR 1.04, 95%CI 1.02-1.06), and disease severity indicator being included via CAG (versus CT scan)(HR 2.80, 95%CI 1.44-5.46) was associated with a higher risk for MACE.

**Figure 1.**
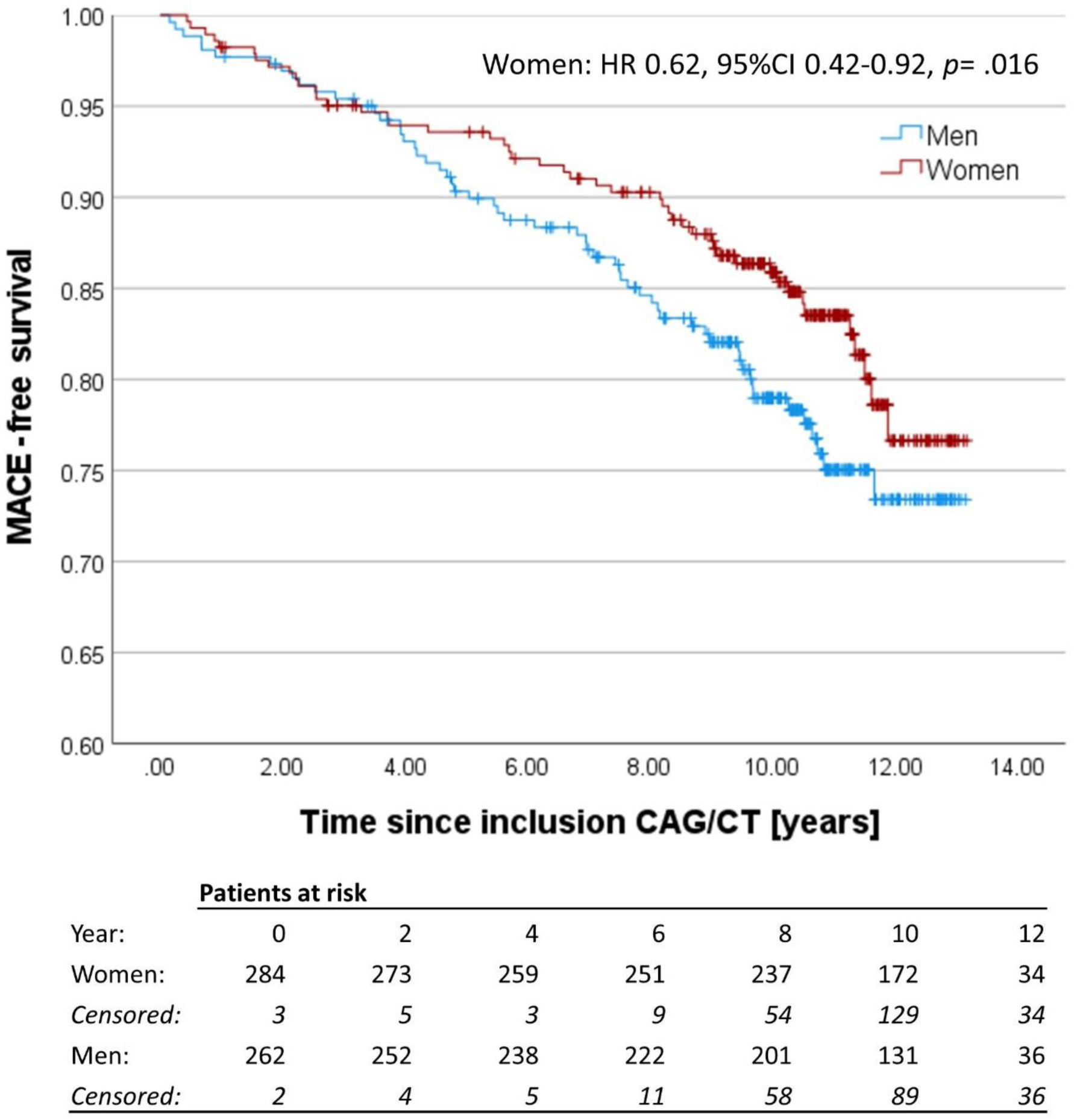
Kaplan-Meier curve of MACE stratified for sex.

Additional analyses were done to examine the risk of the psychosocial factors for MACE stratified by sex (Supplemental Table S1) and moderated by sex (Supplemental Table S2, adjusted for age). Sex stratified findings decreased overall power and rendered more findings nonsignificant. However, the overall pattern is that for women, after adjustment for disease severity and lifestyle factors psychosocial factors were not or no longer significantly associated with MACE. For men, elevated fatigue (HR 1.04, 95%CI 1.00-1.09) and physical limitation (HR 0.99, 95%CI 0.97-1.00) remained significantly associated with MACE. In age adjusted models the psychosocial factors, sex, and their interaction were examined, showing no significant interactions between psychosocial factors and sex (Supplemental Table S2).

The risk of the Type D personality traits with MACE was further explored using both the Z-transformed negative affectivity and social inhibition scores with their quadratic terms in the Cox regression model adjusted for age and sex, showing no significant associations with MACE (Supplemental Table S3). Sensitivity analysis of the ANOCA and INOCA groups compared to ‘other’ NOCAD group adjusted for age and sex showed no significant higher risk for either ANOCA (HR 0.98, 95%CI 0.54-1.80, p=.565) or INOCA (HR 1.19, 95%CI 0.66-2.11, p=.951) with MACE. When excluding non-cardiac mortality (Supplemental Table S4) depressive symptoms according to the BDI, fatigue, and physical limitation were significantly associated with MACE, when adjusted for age and sex, but no longer in models adjusting for disease severity (CAG versus CT) or lifestyle factors. Age was no longer significantly associated with MACE when excluding non-cardiac mortality.

## Discussion

In patients with NOCAD with a median 10 years after index diagnostic CAG or CT scan in total 19% experienced MACE, with a lower risk of adverse outcomes for women compared to men. Covariate adjusted models showed that a lower positive mood, higher fatigue, and lower physical limitation were significantly associated with MACE. This risk seems to be more pronounced in men than in women, though no significant interactions between sex and psychosocial factors were present. Depressive symptoms were predictive of MACE in the age and sex adjusted model, but no longer after further adjustment for confounders including disease severity and lifestyle factors. Noticeable is that Type D personality, anxiety, hostility, mental and physical health status, and angina frequency and stability were not significantly associated with MACE in this patient group. No differences between the INOCA or ANOCA groups emerged, and excluding noncardiac mortality cases rendered more findings nonsignificant.

### Comparability of MACE outcomes

Other studies which examined outcomes in patients with NOCAD or INOCA often included patients with a broader range of NOCAD. Huang and colleagues examined all-cause mortality after 9 years, and included both patients without any visible wall irregularities, as well as patients with either 1, 2, or 3 vessel NOCAD. The AER for all-cause mortality in their 1,2, or 3 vessel disease NOCAD group combined was 1.3% (113 cases/989 patients/9 years, Table 2 findings), with a higher risk for age, men, and 3-vessel disease^22^, which is in line with the findings of our study (1.1%). A higher risk of adverse outcomes for the CAG group compared to the CT group is in line with the meta-analysis of Wang and colleagues^23^. Sedlak and colleagues observed that in the first year after inclusion, women had a higher risk of MACE compared to men, though no sex differences were present from year 1-3^24^. Visual inspection of the Kaplan-Meier curve stratified for sex in the present study shows that sex differences became apparent after four years. Herscovici summarized annual MACE rates in studies, showing an annual MACE rate between 0.9%-2.4%^18^, comparable with the 2% observed in the present study. The WISE study in women with CAG-detected minimal CAD (≥20% but <50% stenosis) reported an all-cause mortality rate of 1.7%, and cardiac mortality was 1.1% AER over 10 years ^25^. Event rates for serious adverse outcomes were recently estimated to be about 3% per year^10^. Notably, each study has slightly different inclusion criteria, as well as different criteria for MACE. A future update of a meta-analysis of risk of adverse outcomes in patients with NOCAD or INOCA including examining sex differences remains to be done.

### Psychosocial factors and health status associated with MACE

Most pronounced findings of the present study were the finding that a lower positive mood, higher fatigue, and lower physical limitation were related to a higher risk of MACE in patients with NOCAD. Depressive symptoms were no longer associated with MACE after adjustment for confounders, which may be due to lack of sufficient statistical power to reliably detect an association. At the same time, there was an absence of anxiety, Type D personality, and hostility with MACE, nor were indicators of health status and angina associated with MACE. Fatigue is a core component of vital exhaustion, which is well observed risk factor for cardiovascular events^8^. In terms of mechanisms, an autonomic imbalance has been hypothesized for the association between exhaustion and cardiovascular risk^26^. Likewise positive psychological wellbeing is related to reduced prognostic cardiovascular events in people with heart disease^27^. The physical limitation scale was a modified version of the SAQ, which in our study represents limitations due to broader defined ‘cardiac problems’ rather than due to ‘chest pain, chest tightness, or angina’. For physical limitation, a poor (0-24 range), fair (25-49 range), and even good (50-74 range) score on the physical limitation scale of the SAQ is associated with mortality^28^. In the present study, the health status subscale ‘physical limitations’ was on average good to excellent, and still predictive of a reduced risk for MACE. These factors show a protective association of experiencing less physical limitations, less fatigue, and better positive mood. This provides a potential starting point for interventions, e.g. aimed at enhancing positive mood^29^, or aimed to develop a greater psychological flexibility rather than addressing negative mental health states, for example by using ‘acceptance and commitment therapy’ (ACT)^30^.

Psychosocial factors, especially depressive symptoms, but also anxiety, Type D personality, and hostility have consistently been associated with MACE^3,31–33^, which is acknowledged in current guidelines^1,2^. A difference between these studies and the present cohort is that these studies predominantly followed patients with obstructive CAD, who had either acute coronary syndrome, a myocardial infarction, or who received invasive treatment such as PCI or CABG. In line with obstructive CAD, those studied populations usually have a majority (70%-80%) of men included. An exception is the WISE cohort study which observed somatic depressive symptoms, but not anxiety by itself to predicted adverse outcomes in women^34^. However further stratification for obstructive versus nonobstructive origin was not examined, nor were other psychosocial factors such as hostility described for outcomes. In the present study, adverse mental health such as anxiety, Type D personality, and hostility were not predictive of MACE, and findings were unclear for depressive symptoms. The differences in findings may be due to several factors: differences in pathophysiological mechanisms in nonobstructive versus obstructive CAD^10^, which are also associated with psychosocial distress^35^, differences in health behavior, age, and sex differences in people with obstructive versus nonobstructive CAD^36^. It is relevant to examine the risk of psychosocial factors for outcomes further stratified for obstructive versus nonobstructive IHD.

### Sex differences in psychosocial factors and outcomes

A consistent secondary finding, in addition to age being a risk for MACE, is that women presented with a lower risk for MACE compared to men. Though no significant sex interaction was observed, sex-stratified findings showed that, after adjustment, only in men, fatigue and physical limitations were associated with MACE. A notable limitation is that power is reduced in these secondary analyses. The absence of an interaction of psychosocial factors with sex, as well as the absence of an association of negative mental health states with MACE is relevant information in terms of gender bias. NOCAD remains underrecognized in general practice^37,38^. Women with NOCAD, more often than men, are often dismissed from specialized care^37^, their symptoms being ascribed to mental health state such as anxiety^39^, and they experience not being taken seriously in their symptom presentation^38^. One of the implications of the present findings is that though psychosocial care could and should be offered for patients to deal with their disease burden^38^, an intervention aimed to reduce depressive symptoms or anxiety is unlikely to affect their prognosis.

### Limitations

A limitation of the present study is that it is a single center cohort study, which could limit the generalizability of the findings towards patients not receiving appropriate care, other countries or ethnic subgroups which are currently underrepresented. Though psychosocial factors were measured at baseline, 12, and 24 months after inclusion, no potential changes over time for the psychosocial factors were taken into account in the present study. The follow-up time is a strength of the present study, though the power in secondary analysis was likely insufficient to reliably detect event risk factors within subgroups. The present study is a combination of patients being included via CAG (70%) as well as CT scan (30%), which created a more heterogeneous group of patients. Moreover, we did not solely focus on patients with ischemia or angina as an inclusion criteria, but included all consecutive patients who had nonobstructive coronary arteries, though no significant differences between these three groups emerged for MACE. A limitation is the absence of more robust data regarding microvascular dysfunction, which was not part of clinical practice between 2009-2013. A limited number of fractional flow reserve (FFR) measurements were performed during CAG, which was upcoming at that time, these were all within normal range.

In conclusion, in patients with CAG or CT scan detected nonobstructive CAD, after a follow-up of 10 years, in total 19% MACE events occurred, with a lower risk for women. A higher positive mood and experiencing less physical limitations were related to a lower risk for MACE, whereas more fatigue was related to a higher MACE risk, without significant sex interactions. No consistent significant risk for depressive symptoms, anxiety, Type D personality, hostility, mental and physical health status, or angina with MACE was observed. Positive mood, fatigue and physical limitations may benefit from specific interventions, and though reducing psychosocial distress is an intervention aim by itself, it is less likely to affect MACE in patients with NOCAD.

## Acknowledgement

We are indebted to the patients who voluntarily contributed to the TWIST study, and who inspired us to initiate subsequent research projects. Since the initiation of this study many students and colleagues have collaborated for short or longer time. Each has been acknowledged or coauthored in a previous research paper. For the present study we would specifically like to acknowledge Liselot Visser, Amber Beltz, Abdeljalil Bel-Elkatib for contributing to collecting outcome information. The ETZ science office, Marjan van den Brink, ETZ cardiology department Karin de Beer, and ETZ IT support Femke van Riel for their support during the data collection.

## Data availability statement

The data underlying this article will be shared on reasonable request to the corresponding author.

## Supplemental Material

Table S1-S3

Figure S1

---

**Supplemental Table S1.** Sex-stratified Cox proportional hazard of psychosocial factors with MACE in patients with NOCAD.

|  | Women<br>Age and sex<br>adjusted | Women<br>Complete<br>adjusted | Men<br>Age and sex adjusted | Men<br>Complete<br>adjusted |
| --- | --- | --- | --- | --- |
| *Psychological distress* | HR (95%CI) | HR (95%CI) | HR (95%CI) | HR (95%CI) |
| Negative Affectivity [z(NA)]<sup>1</sup> | 1.06 (0.76-1.47) | 1.06 (0.77-1.47) | 0.99 (0.72-1.36) | 0.91 (0.65-1.27) |
| Social inhibition [z(SI)]<sup>1</sup> | 1.11 (0.79-1.57) | 0.99 (0.70-1.40) | 1.21 (0.87-1.68) | 1.23 (0.89-1.71) |
| Interaction NA*SI [z(NA)*z(SI)]<sup>1</sup> | 0.87 (0.64-1.19) | 0.89 (0.65-1.20) | 0.99 (0.72-1.35) | 0.99 (0.72-1.36) |
| Type D personality [DS14; dich] | 1.47 (0.81-2.67) | 1.25 (0.68-2.30) | 1.29 (0.73-2.28) | 1.16 (0.65-2.06) |
| Depressive symptoms [BDI total] | 1.02 (0.98-1.05) | 1.01 (0.97-1.04) | **1.04 (1.01-1.08)\*\*** | 1.03 (1.00-1.07)† |
| Cognitive-affective component [BDI cogn]<sup>2</sup> | 0.99 (0.92-1.06) | 0.98 (0.92-1.05) | 1.03 (0.97-1.10) | 1.02 (0.96-1.09) |
| Somatic component [BDI som]<sup>2</sup> | 1.07 (0.97-1.19) | 1.06 (0.95-1.17) | 1.07 (0.96-1.19) | 1.05 (0.94-1.18) |
| Depressive symptoms [HADS-D] | 1.07 (1.00-1.15)† | 1.06 (0.99-1.14) | 1.06 (1.00-1.13)† | 1.04 (0.97-1.11) |
| Anxiety [HADS-A] | 1.05 (0.98-1.12) | 1.04 (0.97-1.11) | 1.03 (0.96-1.09) | 1.01 (0.95-1.07) |
| Positive mood [GMS-PA] | **0.96 (0.92-1.00)\*** | 0.97 (0.93-1.00)† | **0.97 (0.93-1.00)\*** | 0.98 (0.94-1.01) |
| Hostility [CMH] | 0.98 (0.93-1.05) | 0.97 (0.92-1.03) | 1.04 (0.99-1.09) | 1.03 (0.97-1.08) |
| *Health status and Quality of life* |  |  |  |  |
| Fatigue [FAS] | 1.03 (0.99-1.08) | 1.03 (0.98-1.07) | **1.05 (1.02-1.09)\*\*** | **1.04 (1.00-1.09)\*** |
| Physical limitation [modified SAQ] | **0.99 (0.97-1.00)\*** | 0.99 (0.98-1.01) | **0.98 (0.97-0.99)\*\*** | **0.99 (0.97-1.00)\*** |
| Angina stability [modified SAQ] | 1.00 (0.98-1.01) | 1.00 (0.98-1.01) | 1.00 (0.99-1.01) | 1.00 (0.99-1.01) |
| Angina Frequency [modified SAQ] | 0.99 (0.98-1.01) | 0.99 (0.98-1.01) | 1.00 (0.98-1.01) | 1.00 (0.99-1.02) |
| Treatment Satisfaction [modified SAQ] | 1.00 (0.98-1.01) | 1.00 (0.98-1.01) | 0.99 (0.98-1.01) | 0.99 (0.98-1.01) |
| Disease perception/ Quality of life [modified SAQ] | 0.99 (0.97-1.00) † | 0.99 (0.97-1.00) | 0.99 (0.98-1.00)† | 0.99 (0.98-1.01) |
| *SF36 derived PCS and MCS* |  |  |  |  |
| Physical component [PCS] | 0.98 (0.95-1.00) † | 0.98 (0.95-1.01) | **0.98 (0.95-1.00)\*** | 0.98 (0.96-1.01) |
| Mental component [MCS] | 0.99 (0.97-1.02) | 1.00 (0.98-1.02) | 0.98 (0.96-1.00) | 0.99 (0.97-1.01) |
| *Covariates<sup>3</sup>* |  |  |  |  |
| Age at baseline [years] | **1.06 (1.02-1.10)\*\*** | **1.04 (1.01-1.08)\*** | **1.04 (1.01-1.07)\*\*** | **1.04 (1.01-1.07)\*** |
| Inclusion via CAG [versus CT] |  | **3.07 (1.08-8.74)\*** |  | **2.44 (1.02-5.82)\*** |
| Hypertensive [versus not] |  | 6.65 (0.9-48.91)† |  | 1.22 (0.55-2.73) |
| BMI |  | 0.99 (0.92-1.06) |  | 0.99 (0.92-1.07) |
| Physical active [versus inactive] |  | 0.77 (0.41-1.45) |  | 0.82 (0.46-1.46) |

Each psychosocial factor was examined separately with the exception of continuous Type D and BDI subscales
The complete adjusted model included sex, age, disease severity (CAG versus CT group, hypertension), and lifestyle factors (BMI and being physically active)<sup>1</sup> Continuous Type D personality variables were combined
<sup>2</sup> Subscales of BDI; somatic and cognitive/affective component were combined
<sup>3</sup> Covariates taken from model with fatigue based on FAS, but findings are representative for models with other variables.

**Supplemental Table S2.** Hazard ratio’s of psychosocial variables, sex, and their interaction associated with MACE.

|  | Psychosocial variable | Sex (Women) | Interaction of sex with psychosocial variable |
| --- | --- | --- | --- |
| *Psychological distress* | HR (95%CI) | HR (95%CI) | HR (95%CI) |
| Negative Affectivity [z(NA)] | 0.97 (0.71-1.33) | **0.60 (0.39-0.91)\*** | 1.10 (0.70-1.73) |
| Social inhibition [z(SI)] | 1.22 (0.88-1.70) |  | 0.86 (0.54-1.37) |
| Type D personality [DS14; dich] | 1.30 (0.73-2.29) | **0.59 (0.36-0.98)\*** | 1.15 (0.50-2.61) |
| Depressive symptoms [BDI total] | **1.04 (1.01-1.08)\*\*** | 0.75 (0.39-1.44) | 0.98 (0.93-1.02) |
| Cognitive-affective component [BDI cogn] | 1.03 (0.97-1.10) | 0.66 (0.29-1.47) | 0.96 (0.87-1.05) |
| Somatic component [BDI som] | 1.06 (0.96-1.19) |  | 1.01 (0.87-1.17) |
| Depressive symptoms [HADS-D] | 1.06 (0.99-1.12) | 0.55 (0.27-1.09) | 1.02 (0.93-1.11) |
| Anxiety [HADS-A] | 1.03 (0.97-1.09) | 0.50 (0.24-1.06) | 1.02 (0.93-1.12) |
| Positive mood [GMS-PA] | **0.96 (0.93-1.00)\*** | 0.66 (0.23-1.89) | 0.99 (0.95-1.04) |
| Hostility [CMH] | 1.04 (0.98-1.09) | 1.19 (0.41-3.41) | 0.95 (0.88-1.03) |
| *Health status and Quality of life* |  |  |  |
| Fatigue [FAS] | **1.05 (1.02-1.09)\*\*\*** | 0.92 (0.23-3.79) | 0.98 (0.93-1.04) |
| Physical limitation [modified SAQ] | **0.98 (0.97-0.99)\*\*\*** | 0.39 (0.12-1.28) | 1.00 (0.99-1.02) |
| Angina stability [modified SAQ] | 1.00 (0.99-1.01) | 0.65 (0.21-1.98) | 1.00 (0.98-1.02) |
| Angina Frequency [modified SAQ] | 1.00 (0.98-1.01) | 0.97 (0.14-6.86) | 0.99 (0.97-1.02) |
| Treatment Satisfaction [modified SAQ] | 0.99 (0.98-1.01) | 0.41 (0.07-2.31) | 1.00 (0.98-1.03) |
| Disease perception/ Quality of life [modified SAQ] | 0.99 (0.98-1.00) | 0.67 (0.16-2.78) | 1.00 (0.98-1.02) |
| *SF36 derived PCS and MCS* |  |  |  |
| Physical component [PCS] | **0.98 (0.95-1.00)\*** | 0.57 (0.12-2.79) | 1.00 (0.96-1.04) |
| Mental component [MCS] | 0.98 (0.96-1.00) | 0.38 (0.09-1.58) | 1.01 (0.98-1.04) |

All models are adjusted for age.

**Supplemental Table S3.**
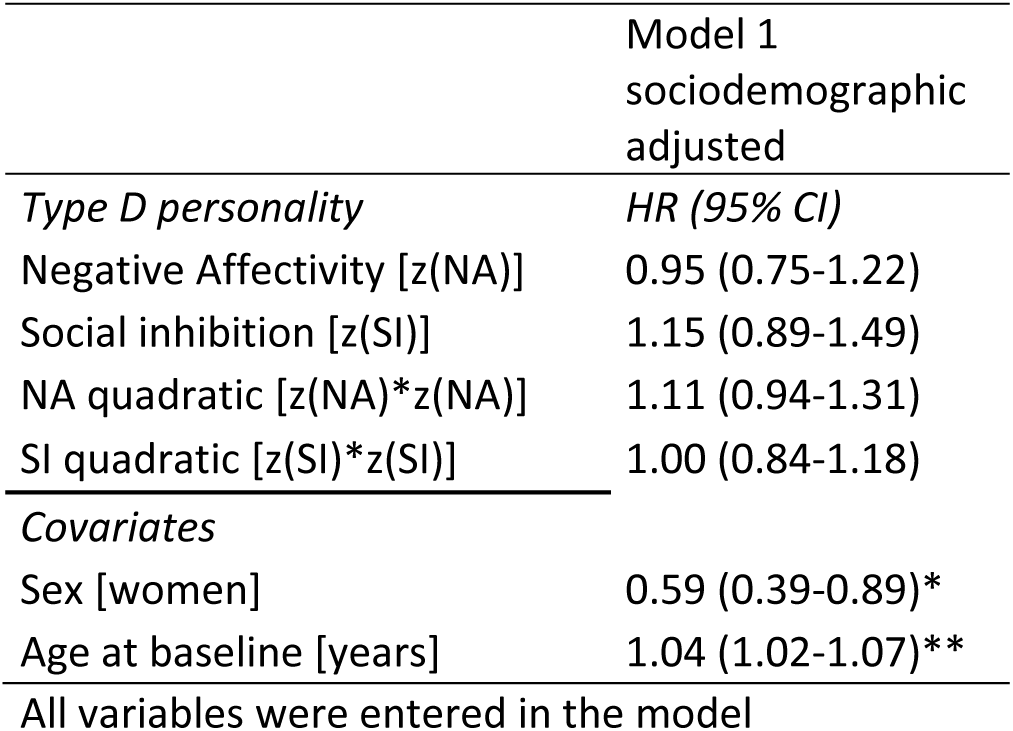
Exploring quadratic terms of Type D personality for MACE.

|  | Model 1<br>sociodemographic<br>adjusted |
| --- | --- |
| *Type D personality* | *HR (95% CI)* |
| Negative Affectivity [z(NA)] | 0.95 (0.75-1.22) |
| Social inhibition [z(SI)] | 1.15 (0.89-1.49) |
| NA quadratic [z(NA)*z(NA)] | 1.11 (0.94-1.31) |
| SI quadratic [z(SI)*z(SI)] | 1.00 (0.84-1.18) |
| *Covariates* |  |
| Sex [women] | 0.59 (0.39-0.89)* |
| Age at baseline [years] | 1.04 (1.02-1.07)** |
| All variables were entered in the model |  |

**Supplemental Table S4.** Cox proportional hazard ratio’s of MACE, excluding non-cardiac mortality.

|  | Age and sex<br>adjusted | Complete<br>adjusted |
| --- | --- | --- |
| *Psychological distress* | HR (95%CI) | HR (95%CI) |
| Negative Affectivity [z(NA)] | 0.95 (0.70-1.29) | 0.90 (0.66-1.23) |
| Social inhibition [z(SI)] | 1.16 (0.87-1.56) | 1.13 (0.84-1.50) |
| Interaction NA*SI [z(NA)*z(SI)] | 0.86 (0.65-1.14) | 0.87 (0.66-1.15) |
| Type D personality [DS14; dich] | 1.15 (0.67-1.97) | 1.03 (0.60-1.78) |
| BDI depressive symptoms [BDI total] | 1.03 (1.00-1.06)* | 1.02 (0.99-1.05) |
| BDI cognitive-affective component | 1.02 (0.96-1.08) | 1.01 (0.95-1.07) |
| BDI somatic component | 1.05 (0.95-1.16) | 1.03 (0.94-1.14) |
| HADS depressive symptoms | 1.05 (1.00-1.12) | 1.03 (0.97-1.10) |
| HADS anxiety | 1.04 (0.98-1.10) | 1.02 (0.96-1.08) |
| Positive mood [GMS-PA] | 0.98 (0.95-1.01) | 1.00 (0.96-1.03) |
| Hostility [CMH] | 1.03 (0.98-1.09) | 1.02 (0.97-1.07) |
| *Health status and Quality of life* |  |  |
| Fatigue [FAS] | 1.04 (1.01-1.08)* | 1.02 (0.99-1.06) |
| Physical limitation [modified SAQ] | 0.98 (0.97-1.00)** | 0.99 (0.98-1.01) |
| Angina stability [modified SAQ] | 1.00 (0.99-1.02) | 1.00 (0.99-1.01) |
| Angina Frequency [modified SAQ] | 0.99 (0.98-1.01) | 1.00 (0.99-1.02) |
| Treatment Satisfaction [modified SAQ] | 0.99 (0.98-1.01) | 0.99 (0.98-1.01) |
| Disease perception/ Quality of life [modified SAQ] | 0.99 (0.98-1.00)† | 0.99 (0.98-1.01) |
| *SF36 derived PCS and MCS* |  |  |
| Physical component [PCS] | 0.98 (0.96-1.01) | 0.99 (0.97-1.02) |
| Mental component [MCS] | 0.99 (0.97-1.01) | 1.00 (0.98-1.02) |
| *Covariates* |  |  |
| Women [versus men] | 0.49 (0.29-0.82)** | 0.54 (0.32-0.91)* |
| Age at baseline [years] | 1.02 (0.99-1.05) | 1.01 (0.98-1.04) |
| Inclusion via CAG [versus CT] |  | 3.00 (1.34-6.70)** |
| Hypertensive [versus not] |  | 2.09 (0.83-5.26) |
| BMI |  | 1.00 (0.93-1.06) |
| Physical active [versus inactive] |  | 0.82 (0.48-1.42) |

N max = 459-478, range between 60-65 events, excluding 67-87 cases due to missing variables in each model.
All psychosocial factors were examined separately, except for the continuous Type D personality variables, and the BDI subscales.
The complete adjusted model included sex, age, disease severity (CAG versus CT group, hypertension), and lifestyle factors (BMI and being physically active)

**Supplemental Figure S1.**
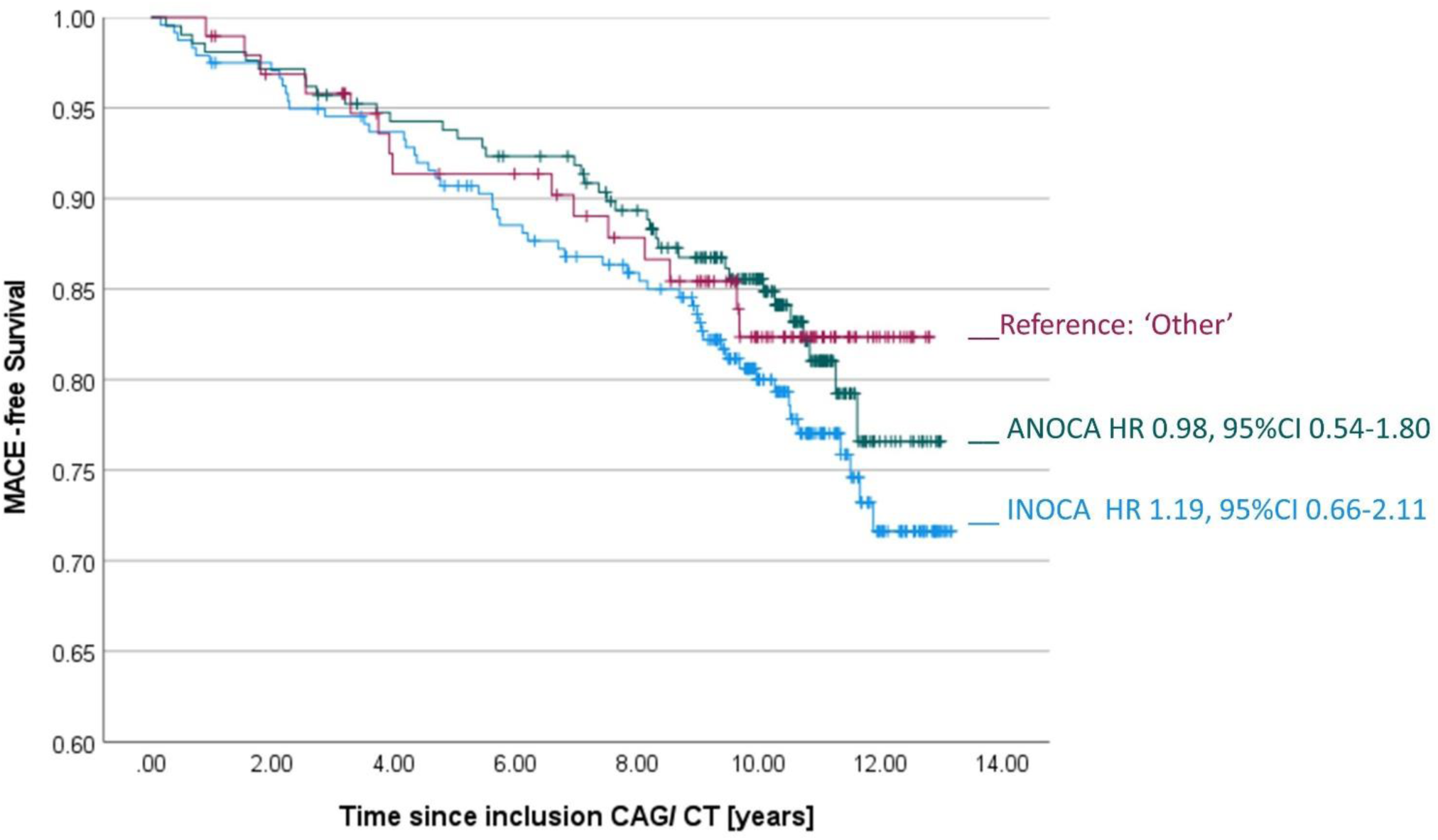
Kaplan-Meier curve for INOCA, ANOCA versus ‘other’ group.

